# Using theory of change to better address social and economic needs in mental health services

**DOI:** 10.1101/2025.08.19.25333973

**Authors:** Helen Baldwin, Anna Greenburgh, Anna Iskander-Reynolds, Dionne Laporte, Hannah Weir, Zara Asif, Mark Bertram, Achille Crawford, Gabrielle Duberry, Shoshana Lauter, Brynmor Lloyd- Evans, Cassandra Lovelock, Aniko Ajozi, Anna-Maria Amato, Pete Hardy, Simon M McKenzie, Guy Swindle, Claire Henderson, Jayati Das-Munshi, Craig Morgan

## Abstract

**Introduction:** Social and economic needs are greater in populations living with mental ill-health compared with the general population. However, services do not currently adequately assess these needs or offer appropriate interventions. A practical roadmap is required to work towards social and economic inclusion as a central component of mental health services.

**Methods:** We used the participatory Theory of Change method to conduct two qualitative workshops with health care professionals, third sector workers, academics and lived experience experts (Workshop 1, n=16; Workshop 2, n=14) in the area served by the South London and Maudsley NHS Trust. We co-developed a Theory of Change model which aimed to outline the key steps needed to put social inclusion at the centre of mental health care services in relation to the largest mental health trust in the United Kingdom (South London and Maudsley), yet with generalisable elements for mental health services in the United Kingdom more broadly.

**Results:** A shared goal for services was developed and agreed by participants of securing “a consistent mental health system that enables individuals to feel loved, valued, and capable of thriving beyond their basic needs”. To progress from the current context to this shared goal, six objective pathways were co-produced to act as a roadmap: (1) Provision of person-centred culturally appropriate care; (2) Advocacy for funding for effective social inclusion services in line with need; (3) Advocacy for funding and support of social and peer programmes; (4) Co-located and localised community-based support hubs; (5) A shared feedback system with social inclusion Key Performance Indicators (KPIs) with an ability to include positive outcomes and drive accountability; and (6) A collaborative community service network.

**Conclusions:** This Theory of Change model offers a tangible framework to put social inclusion at the centre of mental health services. This model can be adapted and translated to other services and settings that are aiming to make social inclusion a core feature of their provision, beyond those in which it was developed.

## 1.0 Introduction

Social and economic needs are substantial and varied among people living with mental ill-health, and particularly for people living with a severe mental illness (1–7), warranting diverse social and economic interventions to address these needs. There is extensive evidence available for some interventions in this field, such as housing and vocational support, with fewer interventions addressing other social and economic domains, such as loneliness, and a notable lack of any evidence in relation to other domains, such as debt, income and social security (8,9). Access to these interventions is not guaranteed and marginalised communities are typically underrepresented in existing provision, despite often experiencing greater need (10,11).

Although social support is vital to good mental health care, and despite the well-evidenced causal role of social and economic adversity in poor mental health outcomes, the existing literature demonstrates that current support is insufficient, and urgent steps are warranted to put social inclusion and economic needs at the centre of mental health care. However, it is not clear what tangible actions are required and, critically, what steps are feasible to make this a reality.

The Theory of Change (ToC) approach is a tool for the co-production of service development. ToC is a framework within implementation science which aims to offer a visual representation of causal pathways through which an intervention (or set of interventions) might achieve their expected impact given resource constraints within the implementation setting (12). This pragmatic approach is particularly fitting for the topic of social inclusion. It allows for multiple causal pathways, levels of interventions and feedback loops (12), better reflecting the complex settings in which social inclusion interventions might be implemented and the intricate interactions between different social and economic needs. This approach also facilitates more thought around the underlying theory for effecting change and so the development of such a framework allows exploration of the tangible steps which would be required to prioritise social inclusion within mental health services.

We therefore aimed to: i) describe how the ToC participatory methodology can be used as a tool for the development and evaluation of social inclusion efforts in mental health services, and ii) present a ToC framework outlining how we can place social inclusion at the centre of mental health care, in collaboration with experts across academia, health and social care, the Voluntary, Community and Social Enterprise (VCSE) sector, policy and lived experience of mental ill-health.

## 2.0 Methods

### Design

We conducted two successive qualitative workshops using the participatory ToC method. Ethics committee approval for this workshop was sought and granted by the King’s College London research ethics committee (Ethics Reference: LRS/DP-23/24-42869). This workshop was co-produced and co-delivered in partnership with members of a project advisory board; this group included health and social care professionals and providers, academics, and lived experience experts. Advisory board members were recruited via multiple channels including the Centre for Society and Mental Health (CSMH) Lived Experience Advisory Board (LEAB) based within King’s College London, South London and Maudsley NHS Foundation Trust and local community organisations with relevant expertise, such as Black Thrive Global.

### Setting

The South London and Maudsley NHS Foundation Trust (SLaM) is the United Kingdom’s (UK) largest mental health trust and one of Europe’s largest mental health providers, serving a population of about 1.3 million people. The catchment area consists of four boroughs/ local government areas, which are among the most deprived in the UK (13), with some specialist services receiving national out-of-area referrals. Service users within SLaM face high levels of economic hardship. An estimated 83% of people in contact with SLaM services between 2005-2020 were in receipt of welfare benefits at some point within this period (14), and approximately 85% of people with severe mental illness in contact with secondary mental health services in SLaM are unemployed (15). Furthermore, there are high levels of disability and poor self-rated health in this population (16) and further social adversities, such as loneliness, are also highly reported in this population (17).

Despite several pioneering initiatives addressing social exclusion of people in contact with SLaM services, these services face high levels of need and are not always routinely signposted to (18). Similarly, our recent service evaluation found that social and economic needs are not always prioritised during assessment (18). This context is not unique to SLaM and similar issues are evident in other catchments within the UK (19), alongside clear gaps in social and economic provision internationally for people living with mental ill-health (8–11).

### Recruitment

We aimed to recruit between 15-20 participants to the workshops on the basis of broad inclusion criteria. Participants all had current or previous experience of delivering, commissioning or accessing mental health care services or associated VCSE organisations in the Trust catchment area or supporting family members to access care.

To facilitate recruitment, members of the project advisory board were invited to attend the workshop and were also asked to recommend relevant experts working or living in South London who would be suitable to attend the workshop, using a word-of-mouth snowballing methodology. Due to the focus on social inclusion in this project, we explicitly considered how best to include the perspectives of communities that typically experience greater social exclusion – this included direct engagement and collaboration with local organisations and charities providing culturally-appropriate care and advocacy. We also circulated recruitment calls via the Lived Experience Advisory Board (LEAB) network within King’s College London.

### Procedures

We adopted a ToC approach to the content and delivery of the workshop (12). An expert facilitator who was proficient in the theoretical framework and practice of ToC was recruited to facilitate the workshop and develop the workshop materials alongside the research team and advisory board members. The workshops were structured according to existing literature detailing best practice for ToC (20,21). Both workshops were held at an external location in South London over one full-day and one half-day respectively. Workshop discussions amongst attendees were minuted manually by the research team and activities were facilitated through the use of posters and post-it notes to create a visual model of change which was modified and refined throughout the day.

Different tools were used to explore the mental health system and ensure that all voices were heard. These included ‘rich pictures’, group discussions, and post-it notes to capture ideas and insights. In the first session, participants explored the mental health system in the trust catchment area, mapping out the key components of the system, the stakeholders involved, key barriers and facilitators to access, and causes and effects of limited social inclusion. Participants also agreed on the conceptualisation of the current challenge which the ToC needed to address, and the ultimate goal to be achieved. The second session then focused on refining the content to develop clear and immediately actionable pathways for change to develop a finalised ToC model.

### Analyses

Based on these discussions and manual notes collected during the first workshop, a first iteration of the ToC model and accompanying documentation was developed to describe each stage and step of the model. Between the workshops, participants were also invited to comment on the first iterations of the ToC model and objective pathways via an interactive interface. These interactive comments, alongside minuted discussions and manual notes from the second workshop, were then used to refine and finalise the ToC model.

## 3.0 Results

### Participants

Forty-seven participants were contacted directly with an invitation to attend the workshop via word-of-mouth, alongside broader invitations which were circulated to two lived experience networks. Of these, 16 people attended the initial ToC development session and 14 attended the follow-up refinement session (one of whom had not attended the first workshop). Participants represented a range of expertise including health and social care professionals and providers, commissioners, academics and lived experience experts across the Trust catchment area. Participants also represented expertise across a range of social life domains, such as welfare and vocation and culturally appropriate care.

### ToC structure

The group defined the current challenge as: “Individuals facing severe mental health challenges are often trapped in a debilitating cycle of poverty which exacerbates their condition. The inefficiencies in the distribution of mental health funding and resources within the system hinder access to adequate care, perpetuating this cycle and preventing recovery and stability.”

The group defined the ultimate goal as: “A consistent mental health system that enables individuals to feel loved, valued and capable of thriving beyond their basic needs.”

Participants then discussed and decided on a number of objective pathways which detailed the steps required to progress from the ‘current challenge’ to the ‘ultimate goal’. Supplementary Figure 1 illustrates this comprehensive theory of change model developed and refined by the workshop participants and clearly presents the steps required to put social and economic inclusion at the centre of mental health services, the impact which can be expected from effectively taking these actions and how each of these actions and impacts may relate and interact across six objective pathways. Due to the large scale of the model, it is presented in full in the Supplementary Materials, whilst each component objective pathway is described in greater detail below.

### Objective pathways

In the first workshop, participants outlined concepts relating to six key objective pathways to create a roadmap from the current challenge to the ultimate goal: (1) Provision of person-centred culturally appropriate care; (2) Advocacy for funding for existing effective social inclusion services in line with need; (3) Advocacy for funding and support of social and peer programmes; (4) Co-located and localised community-based support hubs; (5) A shared feedback system with social inclusion Key Performance Indicators (KPIs) with an ability to include positive outcomes and drive accountability; and (6) A collaborative community service network in the catchment area. Each of these objective pathways is described in detail below (Figure 1-6). Figure 1 illustrates how these objective pathways interconnect to form the full theory of change.

**Figure 1.**
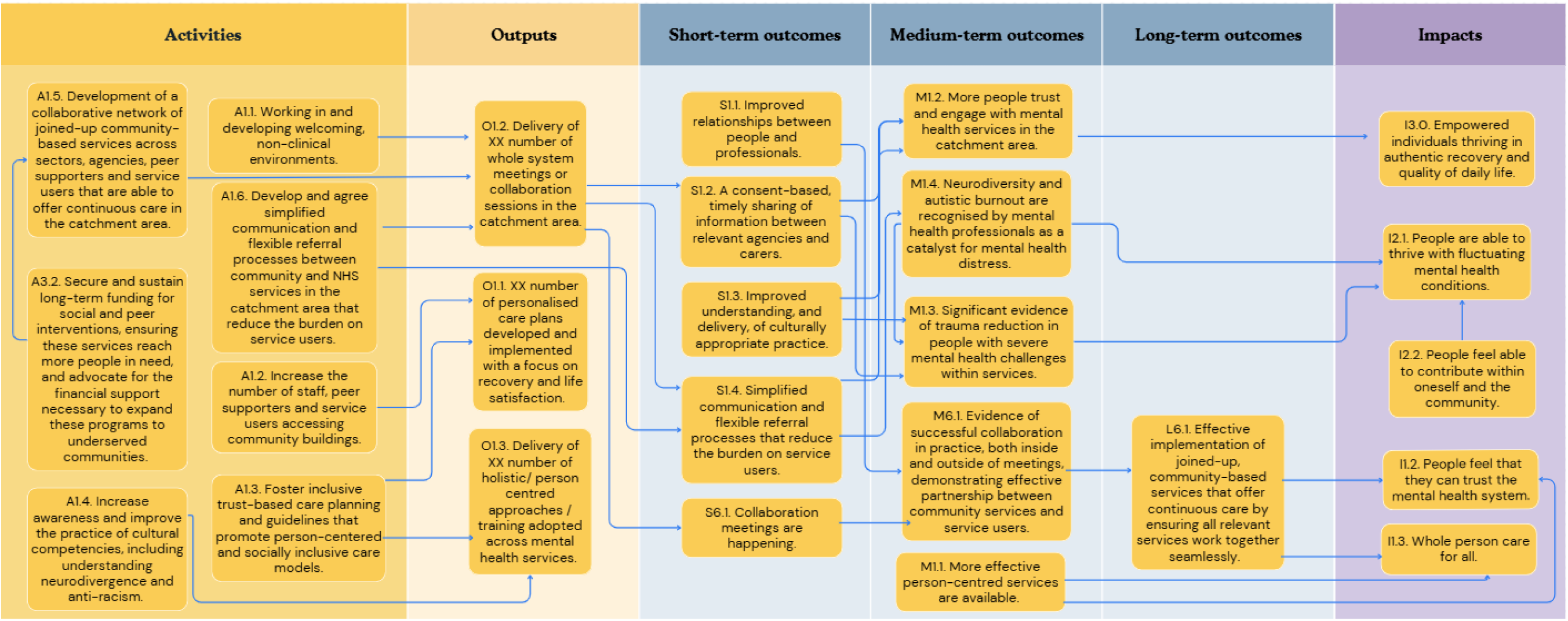
A graphical pathway of the activities, outputs, outcomes and impact of objective pathway 1.0. Here, XX denotes customisable units of output to be tailored to each service using the ToC.

#### 3.1. Provision of person-centred culturally appropriate care

Participants identified person-centred and culturally appropriate care as fundamental to the prioritisation of social inclusion in mental health services. *Figure 2* illustrates this first objective pathway, which was developed and refined throughout the workshops. Key activities required to achieve this goal included: the development of collaborative networks of community organisations and people that are able to offer continuous care and support for social needs in the catchment area; securing sustainable long-term funding for social and economic interventions and peer support interventions to ensure these services reach all who need them - this includes advocating for the financial support necessary to achieve this; the development of simplified referral strategies between the community organisations and services in the catchment area to reduce the burden on the service user to navigate a complex patchwork of services; fostering inclusive Trust-based care planning and guidelines that promote person-centred care; developing welcoming non-clinical environments; increasing the number of staff and peer support workers providing support within community organisations, and in turn, service users who are accessing community organisations; and finally, improving the awareness and practice of cultural competencies and diversity – including neurodiversity and anti-racism.

**Figure 2.**
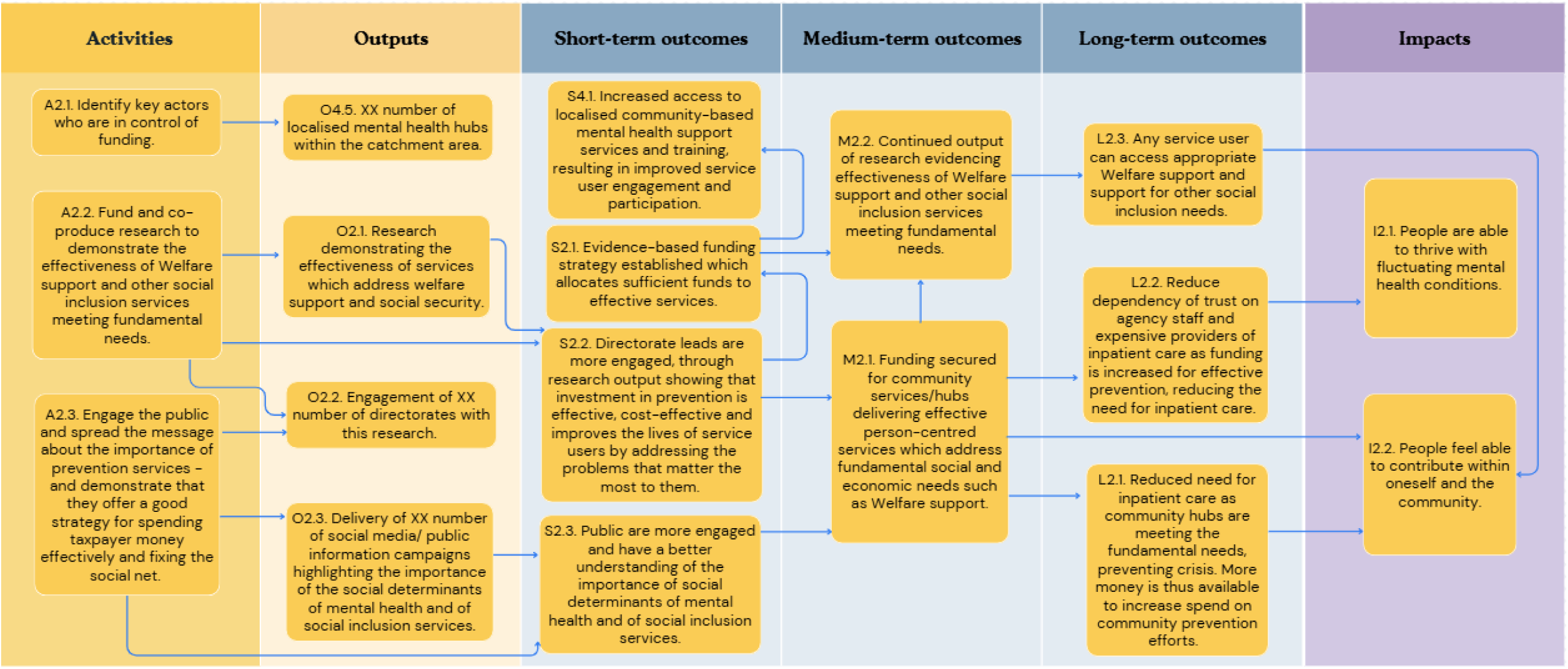
A graphical pathway of the activities, outputs, outcomes and impact of objective pathway objective pathway 2.0. Here, XX denotes customisable units of output to be tailored to each service using the ToC.

#### 3.2. Advocacy for funding for existing effective social inclusion services in line with need

Participants identified consistent funding as an integral meeting the social and economic needs of mental health service users. *Figure 2* illustrates this objective pathway. Participants emphasised the existence of expert teams providing vital support to promote social inclusion; however, that these teams are often overstretched and underfunded, particularly in light of an increasing demand for mental health services and the effects of austerity measures on the health care system. Three key actions were identified: i) identifying the key stakeholders who are in control of funding decisions; ii) the funding and co-production of research to demonstrate the effectiveness and cost-effectiveness of existing welfare support services and other social inclusion services; iii) engaging the public and spreading awareness of preventive care to demonstrate that it is an efficient, cost-effective strategy.

#### 3.3. Advocacy for funding and support of social and peer programmes

Participants identified advocacy for funding and support of social and peer support programmes as another fundamental step needed. *Figure 3* illustrates this objective pathway. Three key actions were identified within this objective pathway: i) as above, the development of a collaborative network of community organisations that are able to offer continuous care; ii) the provision of funding application support to community groups and services; and iii) as above, securing and sustaining long-term funding for social and peer interventions.

**Figure 3.**
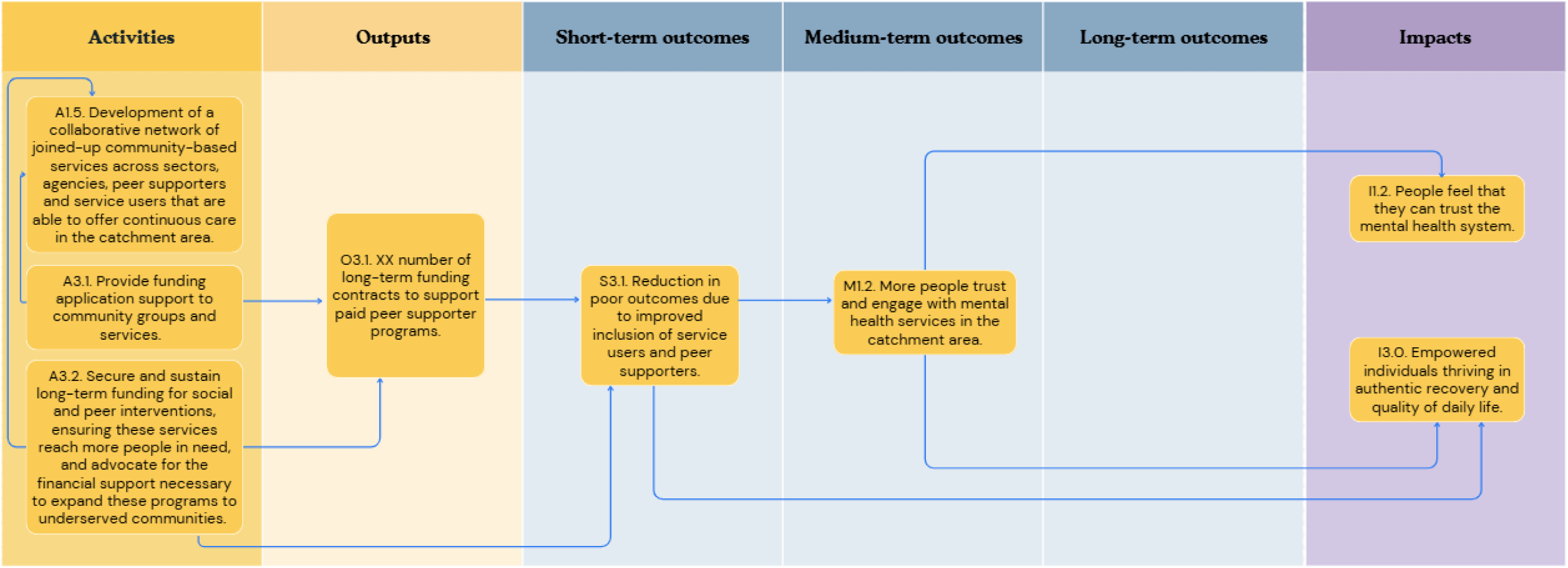
A graphical pathway of the activities, outputs, outcomes and impact of objective pathway objective pathway 3.0. Here, XX denotes customisable units of output to be tailored to each service using the ToC.

#### 3.4. Co-located and localised community-based support hubs

The importance of community-based support hubs was consistently raised by participants. *Figure 4* illustrates this objective pathway. This was highlighted at the same time as advocating for the protection and expansion of specialist distinct services (see *3.2*). Two key actions were outlined here: the expansion and localisation of community-based support hubs, co-located within mental health services, which offer safe spaces for individuals to retrain, learn new skills and receive continuous care; and ii) identifying and expanding upon successful components of the ‘Clubhouse’ model (such as the local ‘Mosaic Clubhouse’ model which has been successful in the catchment area) and adapting this within other boroughs in order to create safe and approachable spaces within communities where ‘Mind and Body’ care and support are readily available.

**Figure 4.**
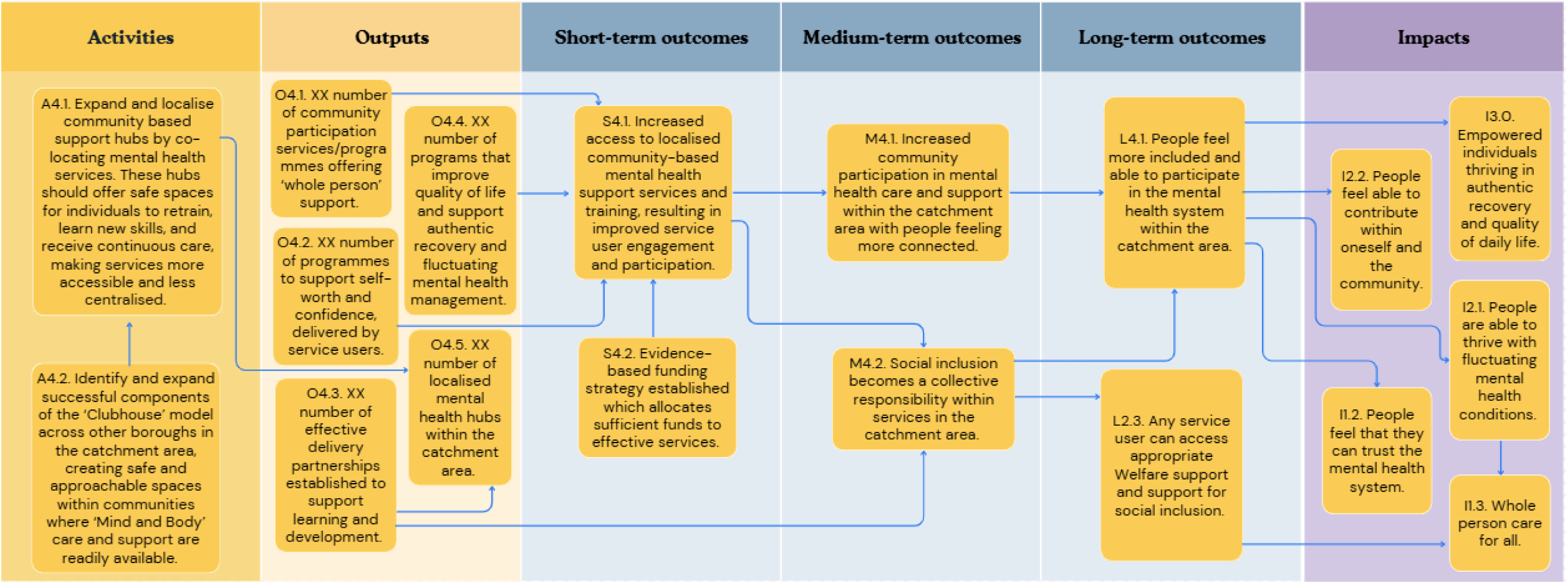
A graphical pathway of the activities, outputs, outcomes and impact of objective pathway 4.0. Here, XX denotes customisable units of output to be tailored to each service using the ToC.

#### 3.5. A shared feedback system with social inclusion Key Performance Indicators (KPIs) with an ability to include positive outcomes and drive accountability

Central to improving social and economic inclusion, participants highlighted the collection of relevant data including a feedback system that allows for the recording of positive events and achievements rather than a solely incident-based reporting system. *Figure 5* illustrates this objective pathway. Five key activities were identified here: i) agreeing on a reasonable timeframe by which to measure KPIs and outcomes – considering enough time to observe reasonable change; ii) Amending the clinical reporting system to better record social inclusion outcomes; iii) Improving person-centred and social inclusion outcome measures; iv) Engaging policymakers to introduce outcome measures and KPIs which include social and personal inclusion, and v) Designing system amendments to be able to measure impact rather than activity.

**Figure 5.**
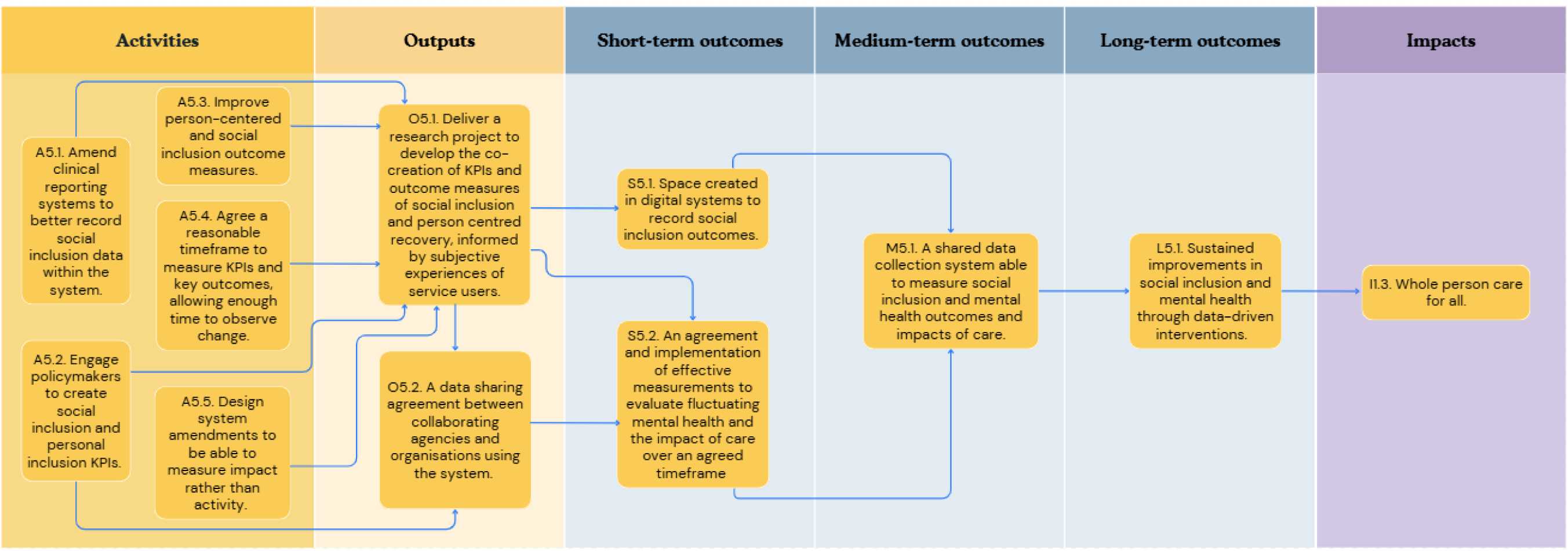
A graphical pathway of the activities, outputs, outcomes and impact of objective pathway objective pathway 5.0.

**Figure 6.**
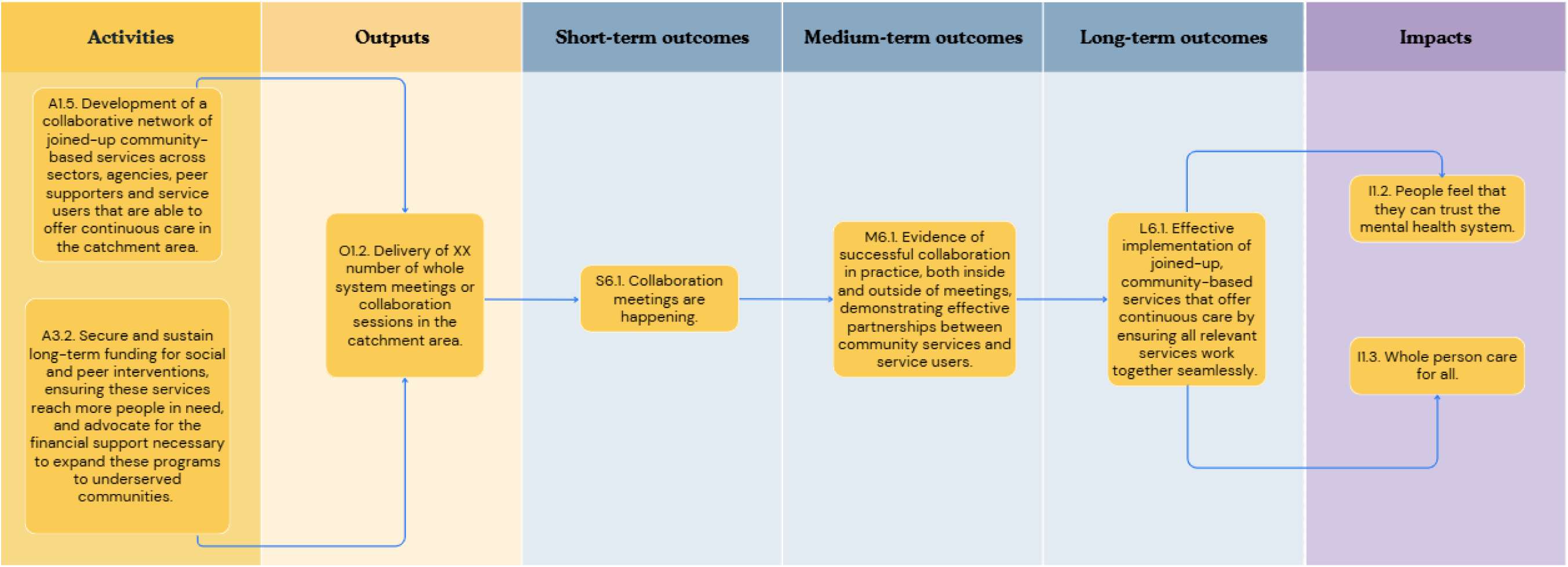
A graphical pathway of the activities, outputs, outcomes and impact of objective pathway objective pathway 6.0. Here, XX denotes customisable units of output to be tailored to each service using the ToC.

#### 3.6. A collaborative community service network in the catchment area

Finally, to support these parallel objective pathways, there is a need for collaboration in the development of networks, alongside the securing and sustaining of funding for social and peer interventions, to create a system which advocates for and achieves a mental health system that offers whole-person care and can be trusted in. *Figure 6* illustrates this final objective pathway.

### Assumptions

Participants highlighted a range of assumptions that underlie the causal pathways of this model – in other words, the necessary factors that must be in place for the model to succeed. These assumptions largely correspond to sociopolitical, organisational, and individual contexts. In terms of sociopolitical contexts, participants assumed that there would be political buy-in to such an approach. In relation to organisational contexts, participants assumed that academic and medical institutions would have the willingness to incorporate key teaching and training on the importance of social and economic inclusion in mental health services; that there would be sufficient funding to retain staff and ensure longevity of staff; that existing expertise will be drawn on in guiding any change; that there are enough trained staff and peer supporters to work with; that effective data sharing processes are in place; that organisations will be open and receptive to this approach; and that peer supporters are paid and supported well in the workplace with opportunities for progression. Finally, in relation to individual-level contexts, participants assumed that there would be underlying staff capacity, motivation and awareness of the importance of social inclusion within health care services and community organisations and to adequately follow care planning and guidleines; that there would be sufficient opportunities for staff and people accessing services to build trust and rapport; there will be representation of people from marginalised and minoritised groups in all aspects of service design, commissioning, management, delivery and peer support; and that individual service users would have the motivation and capacity to engage in their social and economic care needs.

### Indicators

Each ToC outcome was accompanied by a series of indicators which would demonstrate that the outcome had been achieved. For example, for the medium-term outcome ‘Significant evidence of trauma reduction of people with severe mental health challenges within SLaM services’, indicators included: a reduced need for rapid response teams, and a reduction in the use of sedatives, emergent medications, restraints, coercion and other restrictive practice. A full list of indicators is available in the Supplementary Materials.

## 4.0 Discussion

Through collaboration and consultation with a range of experts in the field, we conducted participatory workshops which discussed in depth how we can begin to place social and economic inclusion at the centre of mental health care service delivery within the UK’s largest mental health Trust. The outcome of these workshops was a co-designed ToC model, which maps out the practical steps to move from understanding the current issues to implementing solutions that can make a real impact. The ToC model serves as a shared tool for collaboration, guiding future efforts to improve the social inclusion of individuals with mental health challenges in the Trust catchment area, and beyond. We note that a ToC model is intended for iterative development and flexible use, as such, the ToC coproduced in these workshops does not represent a final, fixed model, but can be built on and adapted where necessary for application in different contexts over time and so can be adapted and applied to other mental health service providers.

Despite the comprehensiveness of the model presented here, there are some limitations to consider in the interpretation of its findings. Whilst a range of expertise informed the development of this ToC model, due to the snowballing method to recruitment, participants did not necessarily represent expertise across the full range of health and social needs and adversities or clinical expertise. Similarly, some of the constituent boroughs of the catchment area (e.g. Lambeth and Southwark) were more represented than others (e.g. Lewisham and Croydon) and so we may have missed important initiatives taking place across the full catchment area or may not have adequately captured the specific barriers and challenges present in these settings.

Furthermore, although lived experience perspectives were integral to the development of this ToC model, there was a greater proportion of health and social care professionals attending the workshops (see Table 1). This may have both underrepresented the perspectives of people with direct experience of accessing, or supporting family members to access, mental health services and may have contributed towards a power dynamic during the workshop activities in which health care professionals demonstrated a more numerate voice. Whilst we did attempt to address these concerns by facilitating activities and discussions in smaller, more balanced groups, this nevertheless speaks to the importance of integrating lived experience in research from conception through to delivery to better identify challenges and solutions to participation in research.

**Table 1.**
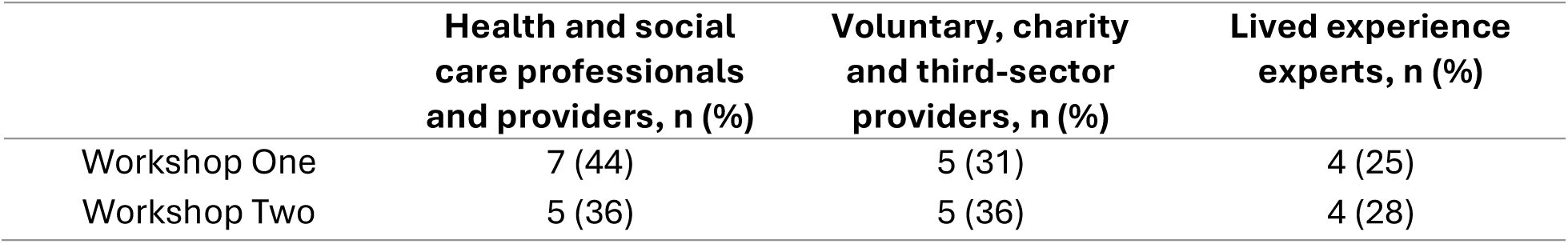
A summary of the workshop participants’ area of expertise.

|  | **Health and social**<br>**care professionals**<br>**and providers, n (%)** | **Voluntary, charity**<br>**and third-sector**<br>**providers, n (%)** | **Lived experience**<br>**experts, n (%)** |
| --- | --- | --- | --- |
| Workshop One | 7 (44) | 5 (31) | 4 (25) |
| Workshop Two | 5 (36) | 5 (36) | 4 (28) |

### Theory of Change

Our ToC provides a practical series of pathways and component steps required to prioritise social and economic inclusion within mental health services. Whilst some of the language and context within this model is specifically developed with the context of South London in mind, the overarching principles of this framework could be amended and applied to a range of other services or settings. Critically, in light of the geographical catchment area and corresponding services which have been developed to serve this population in South London, this model is especially relevant to diverse sociodemographic settings.

Nevertheless, we acknowledge the shifting social and economic contexts within which health care services are operating and thus the responsive need for an adaptable ToC. A core component of the ToC philosophy is that models should be amended and adapted based upon shifting contexts, needs and priorities. Indeed, since these workshops were conducted, NHS-funded, round-the-clock community care pilots have also been established which aim to better provide support for social and economic needs for people living with severe mental illnesses, alongside crisis care, as part of the 10-year Health Plan. Accordingly, we encourage others to utilise the model presented here as a starting point to be built upon. As such, each ToC model may look slightly different for each constituent service. We also note that the limited time of two workshops allowed only for 6 objective pathways to be developed within our Theory of Change, and that, given further resources, many other possible pathways to improve social inclusion might be advanced.

A number of key take-home messages emerged across the workshops. First, the ToC model emphasises the need for person-centred and culturally appropriate care, as opposed to a ‘one-size-fits-all’ approach, which offers more personalised care tailored to possible identities, sociodemographics, and neurodiversity. Indeed, our recent systematic review highlights the potential utility of targeted social and economic interventions and support for marginalised and minoritised communities (10), in parallel with literature reporting the effectiveness of other types of targeted intervention in mental health care (22). Second, participants highlighted the need to better evidence existing services that are addressing social and economic needs, particularly those offering preventive care in the community, to identify the successful elements. This then provides a framework for scaling successful initiatives more widely, allowing for local adaptation to best fit the needs of the target populations. Both of these key themes are underpinned by a crucial need to recognise the vital contributions of VCSE organisations to mental health and social care (23), and to support them to thrive, including advocating for more funding for community and peer-led organisations. A final key theme raised throughout the model is the need for improved reporting and monitoring systems in mental health care which recognise the importance of assessing outcomes relating to socioeconomic inclusion. Indeed, there are already initiatives addressing this, such as the integration of DIALOG+ into routine assessment (24–26).

This process has also highlighted the importance of collaboration in co-designing goals and the means to achieving these goals. Over the course of the workshops, it became apparent that there are clusters of services within the catchment area that are already integral to addressing social and economic needs of the people accessing mental health services. A recent service evaluation from our research group has summarised these efforts in detail (18). We drew on existing knowledge from these experts in the development of this ToC. Further, we involved a range of third sector and voluntary services similarly aiming to address social and economic needs for people experiencing mental ill-health. This highlights the importance of recruiting diverse participants to such participatory methods to more accurately represent the current landscape, including consideration of sectors and specialisms as well as sociodemographic and intersectional features.

Despite the comprehensive range of actions and considerations within the ToC, it is striking that many of the various models of care identified in the most recent systematic reviews of social and economic interventions for people living with mental ill-health (8–11) were not raised during the ToC workshops, most notably, for example, Housing First. This observation may reflect local variations in need compared with the international research context or our local areas of expertise, or conversely may highlight obstacles in the translation of research findings to real-world implementation (27). This calls for a more dynamic partnership between researchers and practitioners to both contribute to current learning and practice to the academic literature and similarly, to better translate research findings into clinical practice alongside recipients and providers of mental health services.

Finally, it is important to reiterate the current overstretched context that this model is intended to operate within. The underlying assumptions listed here that were identified by workshop participants are strong assumptions that are not always met and, equally, may shift in line with the priorities of changing policies and governments and so on. As such, it is important to now assess the feasibility of implementing this framework in practice. Indeed, another key advantage of the ToC methodology is that the listed indicators and impacts lead to an in-built method of performance evaluation by which to assess how well the ToC model is being implemented in practice, and how closely the anticipated impacts are being achieved. Subsequently, if these assumptions are not likely to be met in the current climate, then we must next consider what the key priorities are to take forward which are both considered manageable and capable of enacting real change.

## Conclusions

By design, the ToC participatory approach is an iterative process which should evolve along with changing objectives and contexts. Indeed, the ToC model produced in these workshops can be adapted and utilised by any service aiming to improve their approach to social and economic needs in mental health care. Recommendations from this ToC provide practical next steps, including conducting research projects to evidence existing social inclusion services, and building stronger ties with grassroots organisations and peer supporters. A key takeaway from our workshops is the importance of collaboration – change will only be achieved through continued collaboration with key stakeholders and harnessing the vast wealth of existing knowledge.

## Data Availability

To protect the confidentiality of workshop participants who have opted out of authorship, any raw qualitative data upon which these findings are based will not be made publicly available. All other data produced in the present work are contained in this manuscript.

## Declarations

## Acknowledgements

We would like to thank the participants who attended the theory of change workshops and offered their expertise in the development of this model. This manuscript represents one component of a broader programme of research (the ENRICHED project); we would also like to acknowledge the support offered by Madison Wempe and Katie Chamberlain throughout this project.

## Ethics approval and consent to participate

This study was granted low-risk approval from the King’s College London Research Ethics Committee (Ethics Reference: LRS/DP-23/24-42869), and was conducted in accordance with the Declaration of Helsinki. All participants provided informed consent to participate in the workshops. Some participants also provided consent to public authorship – those who opted out of authorship are not listed on the author line of this manuscript to protect confidentiality.

## Consent for publication

Not applicable.

## Availability of data and materials

To protect the confidentiality of workshop participants who have opted out of authorship, any raw qualitative data upon which these findings are based will not be made publicly available.

