## Supplemental Figure 1 for "Using theory of change to better address social and economic needs in mental health services"

### Theory of Change: A Roadmap for Revitalising SLaM's Mental Health System to Help People with Severe Mental Health Challenges Break Free from Poverty and Thrive in Their Lives.

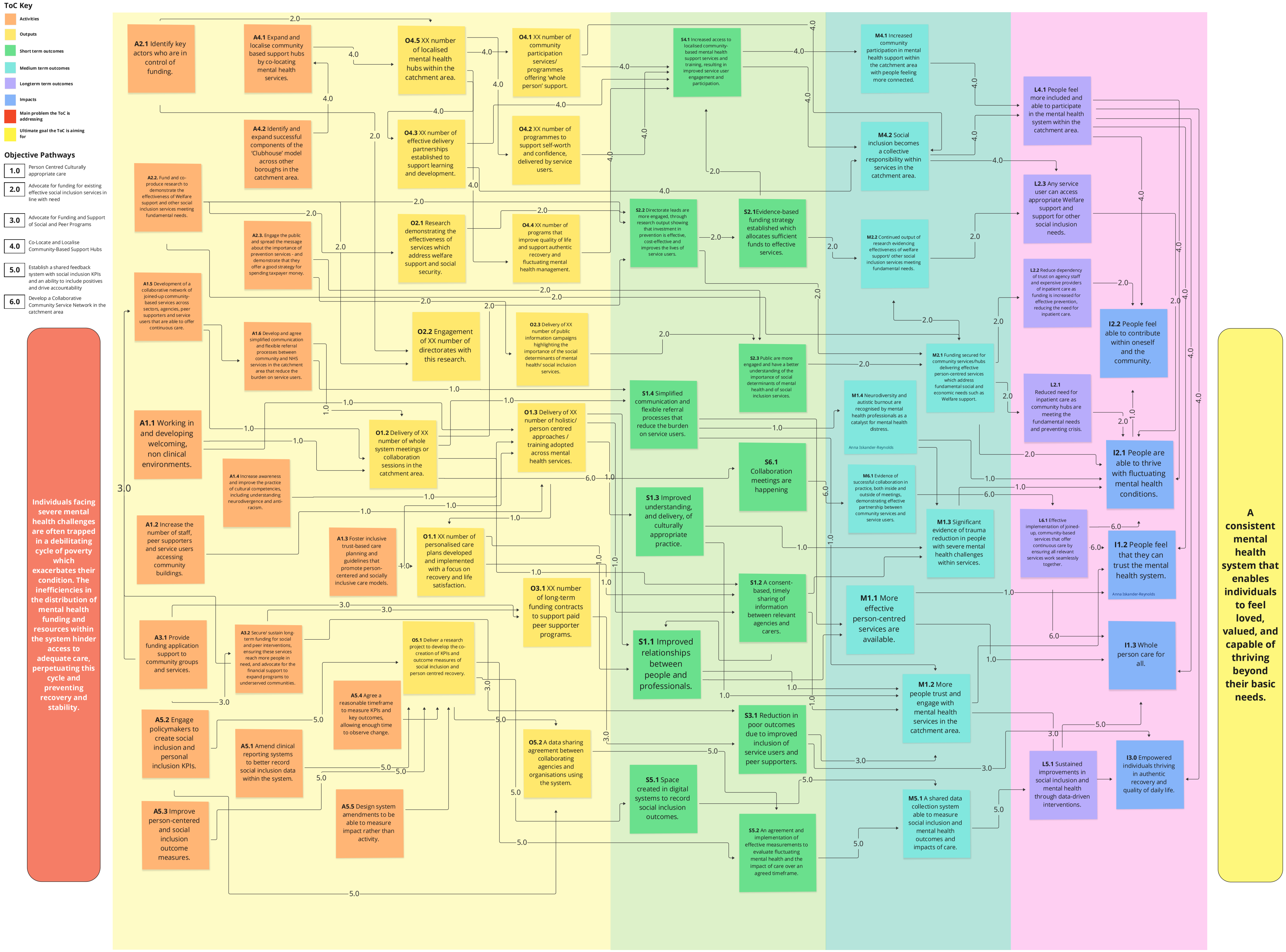
